# Analysis of heterozygous *PRKN* variants and copy number variations in Parkinson’s disease

**DOI:** 10.1101/2020.05.07.20072728

**Authors:** Eric Yu, Uladzislau Rudakou, Lynne Krohn, Kheireddin Mufti, Jennifer A. Ruskey, Farnaz Asayesh, Mehrdad A. Estiar, Dan Spiegelman, Matthew Surface, Stanley Fahn, Cheryl H. Waters, Lior Greenbaum, Alberto J. Espay, Yves Dauvilliers, Nicolas Dupré, Guy A. Rouleau, Sharon Hassin-Baer, Edward A. Fon, Roy N. Alcalay, Ziv Gan-Or

## Abstract

**Background:** Biallelic *PRKN* mutation carriers with Parkinson’s disease (PD) typically have an earlier disease onset, slow disease progression and, often, different neuropathology compared to sporadic PD patients. However, the role of heterozygous *PRKN* variants in the risk of PD is controversial.

**Objectives:** We aimed to examine the association between heterozygous *PRKN* variants, including single nucleotide variants and copy-number variations, and PD.

**Methods:** We fully sequenced *PRKN* in 2,809 PD patients and 3,629 healthy controls, including 1,965 late onset (63.97±7.79 years, 63% men) and 553 early onset PD patients (43.33±6.59 years, 68% men). *PRKN* was sequenced using targeted next-generation sequencing with molecular inversion probes. Copy-number variations were identified using a combination of multiplex ligation-dependent probe amplification and ExomeDepth. To examine whether rare heterozygous single nucleotide variants and copy-number variations in *PRKN* are associated with PD risk and onset, we used optimized sequence kernel association tests and regression models.

**Results:** We did not find any associations between all types of *PRKN* variants and risk of PD. Pathogenic and likely-pathogenic heterozygous single nucleotide variants and copy-number variations were less common among PD patients (1.52%) than among controls (1.8%, false discovery rate-corrected p=0.55). No associations with age at onset and in stratified analyses were found.

**Conclusions:** Heterozygous single nucleotide variants and copy-number variations in *PRKN* are not associated with Parkinson’s disease. Molecular inversion probes allow for rapid and cost-effective detection of all types of *PRKN* variants, which may be useful for pre-trial screening and for clinical and basic science studies specifically targeting *PRKN* patients.

## Introduction

Parkinson’s disease (PD) is a common neurodegenerative disorder with a typical age at onset (AAO) ranging between 60-70 years.^1^ However, a subgroup of patients has early onset PD (EOPD), typically defined as AAO < 50 years.^2^ The most common genetic cause of EOPD are homozygous or compound heterozygous variants in the *PRKN* gene, found in 6.0-12.4% of individuals who present with PD symptoms before the age of 50.^3–5^ *PRKN* has a high rate of single nucleotide variants (SNV) and copy number variations (CNVs), since it is located in a genomic region prone to rearrangements.^6, 7^ *PRKN* encodes Parkin, an E3 ubiquitin protein ligase important in mitophagy.^8^

Neuropathological studies have demonstrated that individuals with biallelic *PRKN* variants diagnosed with PD do not have the typical PD neuropathology, as Lewy bodies are absent in most cases, and the neurodegenerative process is limited to the substantia nigra.^9, 10^ It is therefore possible that patients with biallelic *PRKN* variants represent a distinct subgroup, or arguably a distinct disease with similar clinical features.^10^ Since we are moving towards therapies targeting specific genetic defects in PD (such as *GBA* and *ZRRK2*-targeting therapies), or α-synuclein accumulation^11^ (which is mostly absent in PRKN-related patients),^9^ it is crucial to properly identify these patients. However, the role of rare heterozygous *PRKN* SNVs and CNVs in PD has not been clearly established by association studies,^12^ and it is currently controversial. For example, a previous study with 159 patients and 170 controls showed significant difference in heterozygous *PRKN* SNVs and CNVs between PD patients and controls, while larger studies suggested a lack of association.^13–15^ Additional studies have also shown contradictory results in familial PD, EOPD and late onset PD (LOPD) using SNVs and/or CNVs.^13–34^ Therefore, the role of heterozygous *PRKN* variants remains controversial. Towards future clinical trials targeting *PRKN*, it will be crucial to determine whether heterozygous *PRKN* variants are associated with PD.

To investigate the potential effect of rare heterozygous SNVs and CNVs in PD, we applied a simple, fast and cost-effective method to detect both types of variants. Using targeted next generation sequencing and bioinformatic approaches, we fully sequenced *PRKN* to identify both SNVs and CNVs in a large cohort of PD, including LOPD and EOPD.

## Methods

### Study Population

A total of 2,809 unrelated and consecutively recruited PD patients and 3,629 controls from three cohorts were sequenced, including 1,965 LOPD patients (mean [SD], 63.97±7.79 years, 1,231 men [63%]) and 553 EOPD patients (mean [SD], 43.33±6.59, 374 men [68%]). Age and sex were not available for 291 patients, 88 controls and 22 patients, 4 controls, respectively. After excluding low sequencing quality samples and biallelic *PRKN* carriers, we performed statistical analysis on 6,090 individuals: 2,627 patients and 3,463 controls. The three cohorts are detailed in Table 1 and include: a) a cohort of European ancestry, confirmed by principal component analysis, collected at McGill University, including French-Canadian (mostly recruited through the Quebec Parkinson Network)^35^ and French participants recruited in Quebec, Canada and Montpellier, France b) a cohort recruited at Columbia University, New York, as previously described,^36^ primarily composed of individuals of self-reported European origin and Ashkenazi Jews, and c) a cohort collected at the Sheba Medical Center, Israel, of self-reported Ashkenazi Jewish ancestry, as previously described.^37^ PD was diagnosed by movement disorder specialists according to the UK Brain Bank Criteria, without excluding patients with positive family history ^38^ or the Movement Disorders Society Criteria.^39^ Study protocols were approved by the relevant Institutional Review Boards and all patients signed informed consent before participating in the study.

### Genetic analysis

#### *PRKN* sequencing

All samples were sequenced at McGill University, Canada using the same method. A total of 50 genes were captured using molecular inversion probes (MIPs) and sequenced as previously described.^40^ In brief, probes that specifically target the coding sequences of the genes of interest were designed, followed by capture and PCR amplification of the targeted regions. After adding barcodes, samples were pooled and sequenced at the McGill University and Génome Québec Innovation Centre with Illumina HiSeq 2500/4000. The full protocol is available upon request. Alignment (GRCh37/hg19), quality control and variant calls were done using the Burrows-Wheeler Aligner (BWA),^41^ Genome Analysis Toolkit (GATK v3.8),^42^ and ANNOVAR ^43^ as previously described.^44^ Only rare variants (minor allele frequency, MAF, < 0.01) according to the public database Genome Aggregation Database (GnomAD) ^45^ with a minimum coverage of 30x were included in the analysis. Samples with more than 10% missingness were excluded. The script for these analyses can be found at https://github.com/gan-orlab/MIPVar. We examined all rare exonic variants using the Integrative Genomics Viewer (IGV v 2.7).^46^ All variants were classified using Varsome ^47^ according to the American College of Medical Genetics and Genomics (ACMG) standards and guidelines into five categories: pathogenic, likely pathogenic, uncertain significance, likely benign and benign.

#### Detection and validation of copy number variations

There are four general types of methods to infer CNVs from next-generation sequencing.^48^ Because MIPs target only a small portion of the genome, most CNV breakpoints will not be sequenced. Therefore, only read-depth based methods can be applied for MIPs since other types of methods utilize reads that span breakpoints. In order to detect CNVs, we examined two methods based on read depth for the MIP data, ExomeDepth v1.1.10 ^49^ and panelcn.MOPS v1.4.0 in R.^50^ When using ExomeDepth, each test sample is compared to the best set of reference samples out of 3,629 controls, chosen by the software according to the correlation of the coverage for each probe between the test sample and the reference samples. A filter for samples with correlation above 0.97 per the suggestion of the developer was applied to remove false positives. Panelcn.MOPS also selects the best set of reference samples according to correlation and includes several quality control (QC) steps, such as a minimum user defined depth of coverage per probe. Probes are marked as low quality if their read count shows high variance across the test sample and selected references. To validate CNVs, we performed multiplex ligation-dependent probe amplification (MLPA) using the SALSA MLPA P051-D2 Parkinson probemix 1 kit according to the manufacturer instructions (MRC Holland), which is the gold standard for *PRKN* CNV detection.

#### Quality Control of MIPs for CNV detection

The highest performing parameters were achieved by excluding probes from genes in our library where the average coverage was below 100X in more than 15% of the coding and untranslated regions of the genes. Probes with average coverage below 100X, and samples with average coverage across all genes less than 50X were also excluded. Figure 1 details the numbers of patients and controls in each cohort after different stages of quality control.

**Figure 1.**
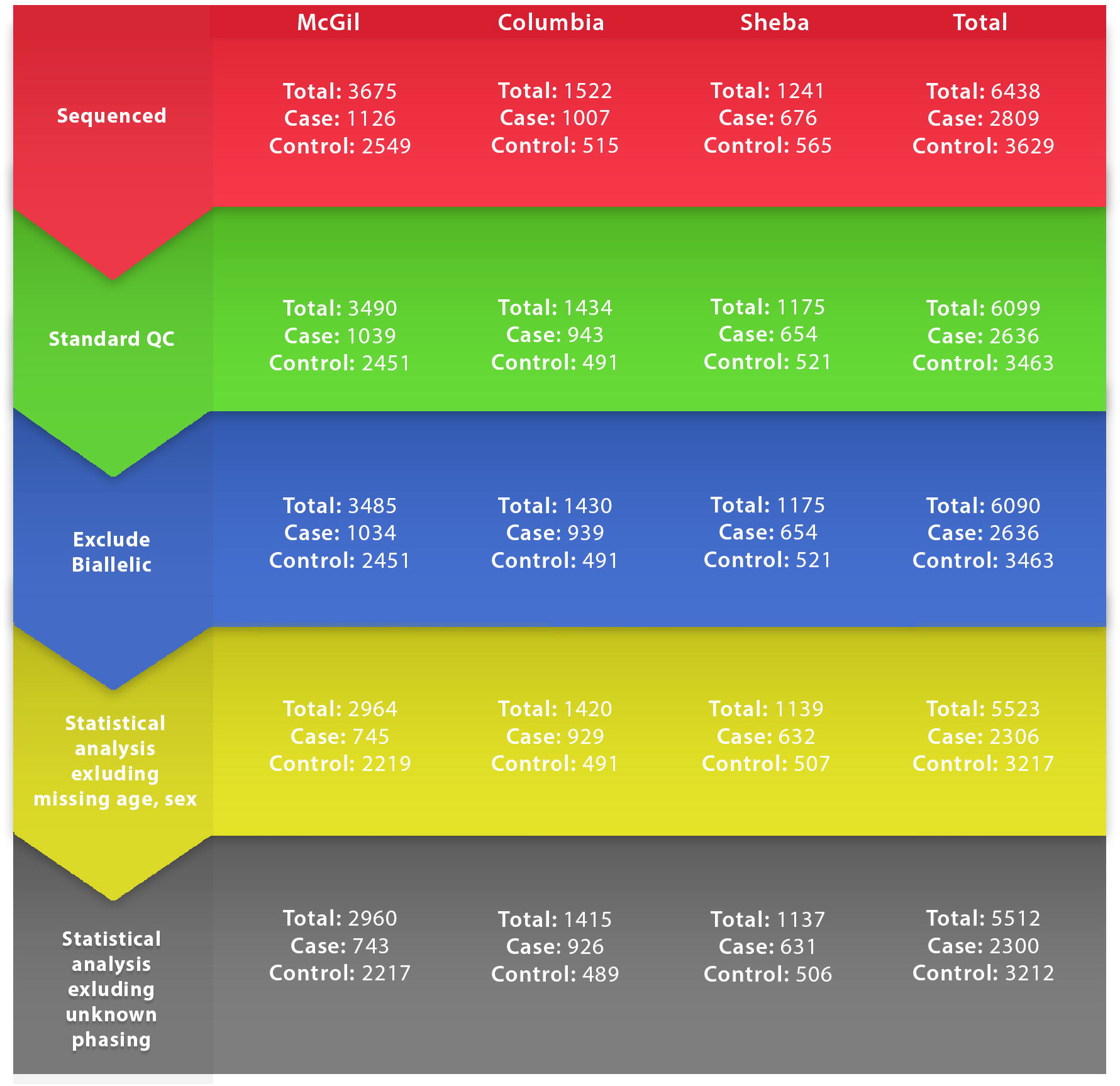
Flow chart of different analysis phases. The flow chart detail the total numbers of patients and controls included in different phases of the analysis. In red, the total number of samples sequenced. In green, the total numbers of samples which passed the quality control phase. In blue, the total numbers of samples after exclusion of 9 patients with biallelic pathogenic and likely pathogenic mutations in *PRKN*. In yellow, the total number of samples included in the analysis aadjusted for age, sex, ethnicity and the presence of *GBA* and *LRRK2*-variants. In grey, the total number of samples included in the analysis after excluding additional samples with potentially pathoigenic biallelic copy number variations that could not be phased, i.e. samples with deletions of consecutive exons, for which we could not determine if they occur on the same allele or if they are biallelic.

#### Statistical Analysis

The associations between rare heterozygous SNVs (MAF < 0.01), heterozygous CNVs and PD were tested using optimized sequence kernel association tests (SKAT-O v1.3.2 in R)^51^ in all cohorts separately, adjusted for age, sex and ancestry as needed. The initial analysis was performed after excluding biallelic carriers of pathogenic and likely pathogenic mutations and adjusting for age, sex, ethnicity and the presence of *GBA* and *LRRK2*variants (Figure 1, yellow).

Rare variants were grouped by: a) CADD score (CADD>12.37), which represent the top 2% of variants predicted to be deleterious, b) functional variants, which include stop gain, nonsynonymous, splice-site and frameshift variants, c) nonsynonymous variants, and d) loss-of-function variants, which include frameshift, splice-site and stop gain variants. A meta-analysis of the results from the three cohorts was performed using MetaSKAT (MetaSKAT v0.80, R)^52^ for heterozygous SNVs, CNVs, and both combined, according to the five ACMG categories (pathogenic, likely pathogenic, uncertain significance, likely benign and benign). Since the age- and sex-adjusted model removes samples without available data on age and sex, we also performed an unadjusted model to avoid this exclusion (Figure 1, blue). We have also repeated all analyses after several additional filtering and adjusting stages, including: adjusting for all *GBA* and *LRRK2* p.Gly2019Ser variant carriers (Supplementary Table 1), excluding these *GBA* and *LRRK2* variant carriers, analyzing only samples with early onset PD (defined as AAO < 50 years), and excluding samples with CNVs in which phasing was not possible (n=8, for example, in a sample with a reported deletion of exons 3-4 the deletion could be on the same allele, or each exon can be deleted on a different allele, Figure 1, grey). The association between heterozygous SNVs, CNVs and AAO of Parkinson’s disease was also calculated using linear regression adjusted for sex and ancestry as needed in all cohorts separately. Here too, patients carrying *GBA* variants or the *LRRK2* p.Gly2019Ser variant (Supplementary Table 1) were excluded and all analyses were repeated. METAL ^53^ was used to performed fixed-effect meta-analysis on all cohorts in the AAO analysis. Since we have performed multiple interdependent analyses, we used a false discovery rate (FDR) correction for multiple comparisons with a FDR-corrected q<0.05 considered as statistically significant.

## Results

### Identification of *PRKN* SNVs and CNVs

The average coverage of *PRKN* (NM_004562) across all samples was 988X, with 98% of nucleotides covered at >30X, and 94% covered at >100X. We identified 199 rare SNVs in 237 patients and 300 controls in the main analysis (Table 2), including nonsynonymous, frameshift deletions and splice site variants in *PRKN* across all cohorts (the specific variants are detailed in Supplementary Table 2).

To identify CNVs, we first aimed to examine which calling method is best suited to properly call CNVs from our MIP targeted sequencing panel. For this purpose, we screened for CNVs in 510 samples using MLPA, the gold standard for CNV detection in *PRKN*. We specifically enriched these samples with EOPD patients to increase the chances to detect CNVs. Out of the 510 samples, 46 carried CNVs in *PRKN* (32 patients and 14 controls). The 32 patients included four homozygous *PRKN* deletion carriers, 17 heterozygous deletion carriers and 11 duplication carriers. Subsequently, we have examined which method (ExomeDepth or panel.cnMOPS) has the highest performance. Except for one deletion for which the MIPs data did not pass QC due to low coverage call rate, deletions and duplications in *PRKN* were identified with 97% sensitivity and 95% specificity using ExomeDepth. In contrast, using the best parameters, panel.cnMOPS had 98% sensitivity but only 54% specificity using samples that passed QC when compared to MLPA. The parameters and CNV call rates for each method are detailed in Supplementary Table 3. Due to its superior performance, we applied ExomeDepth on all cohorts, and identified a total of 62 carriers of CNVs in patients and controls. Supplementary Table 4 details all carriers of CNVs, including heterozygous and bi-allelic carriers of other CNVs or other SNVs.

### Heterozygous *PRKN* SNVs and CNVs are not associated with Parkinson’s disease

To examine the association of rare (MAF < 0.01) heterozygous SNVs and CNVs on risk of PD, we took two approaches. First, we performed a SKAT-O in each cohort to determine whether there is a burden of heterozygous *PRKN* variants of different types. “Pathogenic” variants included pathogenic and likely pathogenic variants, while “non-benign” variants included pathogenic, likely pathogenic and variants of uncertain significance. All CNVs were considered as pathogenic loss-of-function variations. No statistically significant associations were found in any of the SKAT-O analyses (Table 2). Second, we performed a series of meta-analyses by collapsing in each cohort SNVs alone, CNVs alone, and combined. In these analyses too, adjusted for age, sex and ethnicity, no association between heterozygous carriage of *PRKN* mutations and PD was found (Table 2). Pathogenic and likely pathogenic variants were less frequent in patients (1.52%) than in controls (1.8%, *p* = 0.55,, Table 2), suggesting lack of association with risk of PD. In order to avoid the possibility that the exclusion of samples without available data on age and sex had biased the results, we have also performed an unadjusted analysis including all samples. Additional analyses with and without *GBA* and *LRRK2* p.Gly2019Ser variants, with and without CNVs of unknown phasing, and including only samples patients with AAO < 50 have also been performed. In these analyses too, there were no statistically significant differences between patients and controls (Supplementary Table 5-6).

### Heterozygous *PRKN* SNVs and CNVs are not associated with AAO of Parkinson’s disease

The association between rare heterozygous SNVs and CNVs on AAO of PD was examined using linear regression in each cohort alone on the same groups of mutations mentioned in the previous association study. After adjusting for sex, ancestry, and the presence of *GBA* and *LRRK2* variants, we found no association in any analyses. We also performed meta-analysis by collapsing each cohort which yielded no statistically significant results (Table 3). When examining CNVs, the meta-analysis shows an earlier AAO in heterozygous *PRKN* CNV carriers (3.6 years younger compared to non-carriers), but the association was not statistically significant after correction for multiple comparisons. This difference in AAO was mainly driven by an effect of CNVs in the Columbia cohort, which was almost 8 years younger in carriers of CNVs (average AAO of 51.85 years) compared to non-carriers of CNVs (59.44 years). This difference was not statistically significant after correction for multiple comparisons as well. Larger studies for AAO of heterozygous *PRKN* carriers are needed to further study these findings. Association analyses between different types of heterozygous *PRKN* variants and AAO of PD, including with and without *LRRK2* and *GBA* variant carriers, with and without ambiguous phasing (see methods), and in AAO < 50 can be found in Supplementary Tables 7-8. In all analyses, there were no statistically significant associations.

### Identification of *PRKN*-associated parkinsonism patients

Overall, we were able to identify 9 patients with pathogenic or likely pathogenic homozygous and compound heterozygous *PRKN* SNVs and/or CNVs (Table 4). The most common pathogenic SNV in our cohort was p.Gln34ArgfsTer5 mutation, found in 3 (33%) *PRKN* patients, and the most common CNV was heterozygous deletion of exon 3, found in 7 (77%) *PRKN* patients. The average AAO of PD in biallelic *PRKN* SNV/CNV carriers was 28.0 ±7.82 years old.

## Discussion

In the current study, we found that the frequencies of heterozygous SNVs and CNVs in *PRKN* are similar in PD patients and controls. These results do not support a role for heterozygous *PRKN* variants in the risk of PD or its AAO. Of note, in one cohort (Columbia), the average AAO of CNV carriers was about 8 years younger compared to non-carriers (Table 3), yet in the other cohorts there was no difference between CNV carriers and non-carriers. Additional studies on AAO in heterozygous *PRKN* carriers are required to conclusively determine whether or not they are associated with earlier AAO. Since the *PRKN* region is prone to genetic variance,^6^ including multiple SNVs and CNVs, properly genotyping all types of *PRKN* variants could be challenging. Using a simple, fast and cost-effective method, we were able to successfully detect all CNVs, SNVs and indels. With MIPs, deep coverage can be achieved, and the probes always target the exact same region, as opposed to whole-exome or whole-genome sequencing where there is no full overlap between all the reads. When the coverage is high, it provides an advantage that allows for more accurate calls of CNVs as well as SNVs and indels. Using this approach, we have identified 199 rare *PRKN* variants and 62 participants with *PRKN* CNVs, with very high sensitivity and specificity (97% and 95%, respectively, when compared to the gold standard MLPA method). Our approach can therefore be used for large-scale screening of PD cohorts, with only validation of detected *PRKN* CNVs with MLPA, instead of fully screening all patients with MLPA. Of note, we identified 9 patients with pathogenic and likely pathogenic biallelic *PRKN* variants. This number of patients is lower than previously reported in EOPD. It is possible that in Ashkenazi Jewish Parkinson’s disease patients (comprising the entire Sheba cohort and a large portion of the Columbia cohort), the frequency of *PRKN* variants is lower, as evident by the lack of such patients in the Sheba cohort. This is also supported by the Columbia cohort, in which all biallelic *PRKN* patients are of European ancestry and none among the Ashkenazi Jewish origin.

There have been multiple studies analyzing the role of heterozygous *PRKN* mutations with conflicting results, shown in Supplementary Table 9. These conflicts may arise from different screening approaches. Some studies first sequenced all patients for rare SNVs and/or CNVs, then sequenced only for selected variants in controls. This approach will create a bias, as the controls may carry other pathogenic *PRKN* variants. Other studies sequenced all patients and controls for heterozygous SNVs and/or CNVs more systematically, and the majority of them were negative. Systematic analysis, as was done in the current study, will avoid misrepresenting the genetic landscape of the study population. Our results do not support an association between heterozygous SNVs and CNVs in *PRKN* and PD, which is supported by other systematic studies of *PRKN* as shown in Supplementary Table 9.^14–17, 24, 33^ These results also emphasize the need for determining the pathogenicity of different *PRKN* variants, as many variants are currently defined as variants of unknown significance. Having a reliable assay for Parkin activity, as previously suggested, would provide an experimental way to assess pathogenicity of *PRKN* variants.^54^

To further study the potential effect of heterozygous *PRKN* variants, previous studies have compared the rate of 18F-dopa uptake in biallelic *PRKN* patients, asymptomatic heterozygous *PRKN* mutation carriers and healthy controls.^55, 56^ These studies have suggested that some *PRKN* heterozygous carriers may have reduced uptake of ^18^F - dopa, especially in the caudate and putamen. A follow-up longitudinal study by one of these groups, however, suggested that this reduction is subclinical, and that the rate of progression is very slow and unlikely to lead to clinical parkinsonism manifestations.^57^

In recent years, treatments that target specific genes and proteins implicated by human genetic studies, such as *SNCA* (α-synuclein), *GBA* and *LRRK2*, are being tested in clinical trials.^58^ Therefore, identifying patients that may benefit from these trials, or conversely, patients that are less likely to benefit, is crucial. Neuropathological studies on brains of patients with PRKN-associated parkinsonism have demonstrated that the vast majority of patients with biallelic *PRKN* mutations do not have accumulation of α-synuclein and the typical Lewy bodies that are seen in PD.^59^ Since α-synuclein does not accumulate, it is likely that treatment targeting α-synuclein will not be efficient for these patients, who should therefore be excluded from these clinical trials. Furthermore, the neurodegenerative process in PRKN-associated Parkinsonism is limited to the substantia nigra and locus coeruleus, and does not spread to other brain regions.^60^ Since we did not detect an association between heterozygous *PRKN* variants and PD, we recommend that heterozygous carriers of *PRKN* variants should not be excluded from such trials, as it is likely that the presence of heterozygous *PRKN* variants in PD patients is due to chance. Clinically, patients with PRKN-associated Parkinsonism are also different, as they have early onset disease, slowly progressing and typically without or with very limited non-motor symptoms.^59^ Therefore, it is important to identify these patients, and our method for rapid and cost-effective detection of *PRKN* variants would be useful for pre-trial screening and for clinical and basic science studies specifically targeting *PRKN* patients.

Although this study examined heterozygous mutations systematically, there are several limitations. The error rate of ExomeDepth CNV detection could affect the results of the association study because not all samples were analysed using MLPA. Furthermore, potentially pathogenic intronic variants have not been examined since intronic regions were not sequenced. In addition, our cohorts were not matched for age and sex. Our controls are on average younger and our patients are predominantly composed of men, yet age and sex were adjusted for when possible. The missing age at onset of patients underpowers our AAO study, however, because data were missing at random, its effect on our results is likely minimal. Another limitation is that in a case-control set-up, phasing cannot be performed, and patients with two variants are considered as compound heterozygous carriers. Since all patients with two mutations had AAO<50, it is likely that indeed they are all compound heterozygous, but we cannot rule out that they carry two variants on the same allele. In addition, individuals with CNVs in consecutive exons are considered as heterozygous carriers, while in fact they can have separate deletions of each exon in different alleles. To examine whether inclusion of these patients affected the results, we repeated the analysis after excluding them, which did not substantially change the results (Supplementary Tables 5-6). An additional limitation of our study is that it includes predominantly individuals of European and Ashkenazi Jewish ancestries. While we adjusted for ancestry in the analysis, studies in additional ancestries are required to determine if heterozygous *PRKN* variants may have a role in PD in other populations.

To conclude, our findings do not support a role for heterozygous *PRKN* variants in PD, and additional large-scale studies are required for a definite conclusion. Our study and the methods we have used provide a framework and a cost-effective method for rapidly screening for all types of *PRKN* variants, which will be useful in future genetic and clinical studies, and for stratification or patient selection for clinical trials.

## Data Availability

Data Availability Statement
The data that support the findings of this study are available from the corresponding author upon reasonable request.

## Acknowledgment

We thank the participants for contributing to the study. GAR holds a Canada Research Chair in Genetics of the Nervous System and the Wilder Penfield Chair in Neurosciences. EAF is supported by a Foundation Grant from the Canadian Institutes of Health Research (FDN grant – 154301). ZGO is supported by the Fonds de recherche du Québec - Santé (FRQS) Chercheurs-boursiers award, in collaboration with Parkinson Quebec, and by the Young Investigator Award by Parkinson Canada. The access to part of the participants for this research has been made possible thanks to the Quebec Parkinson’s Network (http://rpq-qpn.ca/en/). We thank Daniel Rochefort, Hélène Catoire, Clotilde Degroot and Vessela Zaharieva for their assistance.

## Authors’ Roles

1) Research project: A. Conception (EY, ZGO), B. Organization (EY, LK, JAR, FA, MS, SF, CHW, LG, AJE, YD, ND, GAR, SH-B, EAF, RNA, ZGO), C. Execution (EY, UR, LK, KM, JAR, FA, DS).

2) Data Generation: A. Experimental (EY, UR, LK, KM, JAR, FA, MAE, DS, MS, SF, CHW, LG, AJE, YD, ND, GAR, SH-B, EAF, RNA, ZGO).

3) Statistical Analysis: A. Design (EY, ZGO), B. Execution (EY).

4) Manuscript Preparation: A. Writing of the first draft (EY, ZGO), B. Review and Critique (EY, UR, LK, KM, JAR, FA, MAE, DS, MS, SF, CHW, LG, AJE, YD, ND, GAR, SH-B, EAF, RNA, ZGO).

## Funding agencies

This study was financially supported by grants from the Michael J. Fox Foundation, the Canadian Consortium on Neurodegeneration in Aging (CCNA), the Canada First Research Excellence Fund (CFREF), awarded to McGill University for the Healthy Brains for Healthy Lives initiative (HBHL), and Parkinson Canada. The Columbia University cohort is supported by the Parkinson’s Foundation, the National Institutes of Health (K02NS080915, and UL1 TR000040) and the Brookdale Foundation.

